# The global distribution of COVID-19 vaccine: The role of macro-socioeconomics measures

**DOI:** 10.1101/2021.02.09.21251436

**Authors:** Ali Roghani, Samin Panahi

## Abstract

Since coronavirus disease 2019 (covid-19) has continued to spread globally, many countries have progressed clinical trials and started vaccinations at the end of December 2020. This report aims to analyze the association of COVID-19 vaccine distribution and two macro-socioeconomics measures, including human development index (HDI) and Gross domestic product (GDP), among 25 countries till the first week of February 2021. Our results indicate that a higher GDP per Capita is positively associated with higher COVID-19 vaccine distribution. In addition, the result shows HDI does not have a significant relationship with vaccine distribution. Although these macro-socioeconomic measures may be counted as a vital indicator for vaccine distribution, other factors may play roles in vaccine distribution, including well-developed health infrastructure, a centralized political system, and population size.

## Introduction

The world health organization (WHO) announced the COVID-19 global pandemic in the middle of March 2020. The coronavirus outbreak has spread to more than 200 countries globally and has negatively affected the health and economic factors globally. According to the Bureau of an economic analysis report, the world GDP fell by 4.6% in 2020. However, the recovery is expected if there is no delay in vaccine deployment in the world. The WHO has estimated that at least 65% of the given population should be vaccinated to stop the spread of the virus for the population’s immunity. According to a recent article published in Nature news, the leading vaccine developers estimated making sufficient doses of vaccine for approximately one-third of the world population by the end of 2021, which most of this capacity was already pre-ordered by developed rich countries. Therefore, lower-income countries may have to be waitlisted until 2023 (4). The remarkable power of rich countries for purchasing vaccines and health for their people has also been proved by previous studies showing the link between health and wealth for decades, using various methods and evidence for causality in both directions. Rich countries have more resources to provide vaccination for their citizens by medical infrastructure and security to facilitate a mass vaccination program (1). Therefore, a well-developed toolkit can help our understanding regarding long-term economic growth and the value of health interventions. Research has used measures of Gross Domestic Product (GDP) and Human Development Index (HDI) to examine the link between macro-socioeconomic measures and broad health measures (5). One of the most substantial effects of high GDP growth is a more significant capital investment in improving vaccination rates by having great physical and human capital investments (Gutierrez, 2017). In addition, HDI is a statistic composite index of life expectancy, education, and per capita income indicators applied to assess countries’ social health determinants (3). This study examines the relationship between GDP per capita, HDI, and the distribution of COVID-19 vaccine among 25 countries data as of February 5, 2021, to understand how these macroeconomics measures influence people’s vaccination among low to high-income countries.

### Data, Measure, and Method

Data for this analysis comes from Coronavirus Pandemic (COVID-19) data sets of Our Data in World (OIWD)(https://ourworldindata.org/coronavirus).

This data provides daily rates of vaccinations in the country levels. GDP per capita is estimated by calculating all US $ and HDI values, including life expectation, education, and living standard measures. Other measures are used to control the regression analysis, including the number of hospital beds and countries’ population density. Lastly, the outcome is estimated people vaccinated per hundred. Twenty-five countries are included in this study that mostly from high-income countries.

## Results

Table 1 indicates the summary of measures that are used in this study. Indonesia has the lowest GDP per Capita in the analysis. Most of the European countries have a range of $30,000 to $45,0000 GDP per Capita. However, Norway and Luxemburg stand on the top among European countries with $64,800 and $94.277 GDP per Capita. Among Asian countries, United Arab Emirates has the highest GDP per capita. HD’s Range is 0.936 (Germany) and 0.69 (Indonesia).

**Table 1.** Summary of the Measures

| Country | GDP per Capita | Human Development Index | People Vaccinated per Hundred |
| --- | --- | --- | --- |
| Austria | 45436.69 | 0.908 | 2.07 |
| Belgium | 42658.58 | 0.916 | 2.60 |
| Bulgaria | 18563.31 | 0.813 | 0.42 |
| Czechia | 32605.91 | 0.888 | 2.22 |
| Denmark | 46682.51 | 0.929 | 3.28 |
| Estonia | 29481.25 | 0.871 | 2.12 |
| France | 38605.67 | 0.901 | 2.27 |
| Germany | 45229.25 | 0.936 | 2.38 |
| Hungary | 26777.56 | 0.838 | 2.52 |
| Indonesia | 11188.74 | 0.694 | 0.20 |
| Israel | 33132.32 | 0.903 | 36.62 |
| Italy | 35220.08 | 0.880 | 2.25 |
| Lithuania | 29524.26 | 0.858 | 2.68 |
| Luxembourg | 94277.96 | 0.904 | 1.81 |
| Mexico | 17336.47 | 0.774 | 0.48 |
| Norway | 64800.06 | 0.953 | 1.98 |
| Poland | 27216.44 | 0.865 | 2.64 |
| Portugal | 27936.90 | 0.847 | 2.65 |
| Romania | 23313.20 | 0.811 | 3.11 |
| Slovakia | 30155.15 | 0.855 | 2.61 |
| Slovenia | 31400.84 | 0.896 | 2.57 |
| Spain | 34272.36 | 0.891 | 2.68 |
| United Arab Emirates | 67293.48 | 0.863 | 32.26 |
| United Kingdom | 39753.24 | 0.922 | 14.21 |
| United States | 54225.45 | 0.924 | 7.78 |

Figure 1 is the correlation table, indicating a positive relationship between GDP per Capita, HDI, and the percentage of people vaccinated. Both of these correlations are not high; however, GDP has a higher correlation than HDI (0.23 Vs. 0.17). Lastly, this table shows a high positive relationship between GDP per Capita and HDI among the countries in the analysis (0.66). Figure 2 shows the relationship between GDP per capita and people vaccinated per hundred is a linear relationship; however, some countries. have different performances. Indonesia and Bulgaria, with low GDP, have a low vaccination distribution. Most of the European countries and the United States followed the linear relationship of GDP and vaccination distribution. However, with very high GDP per capita, Norway and Luxembourg have a similar performance to other European countries such as Germany and Denmark.

**Figure 1.**
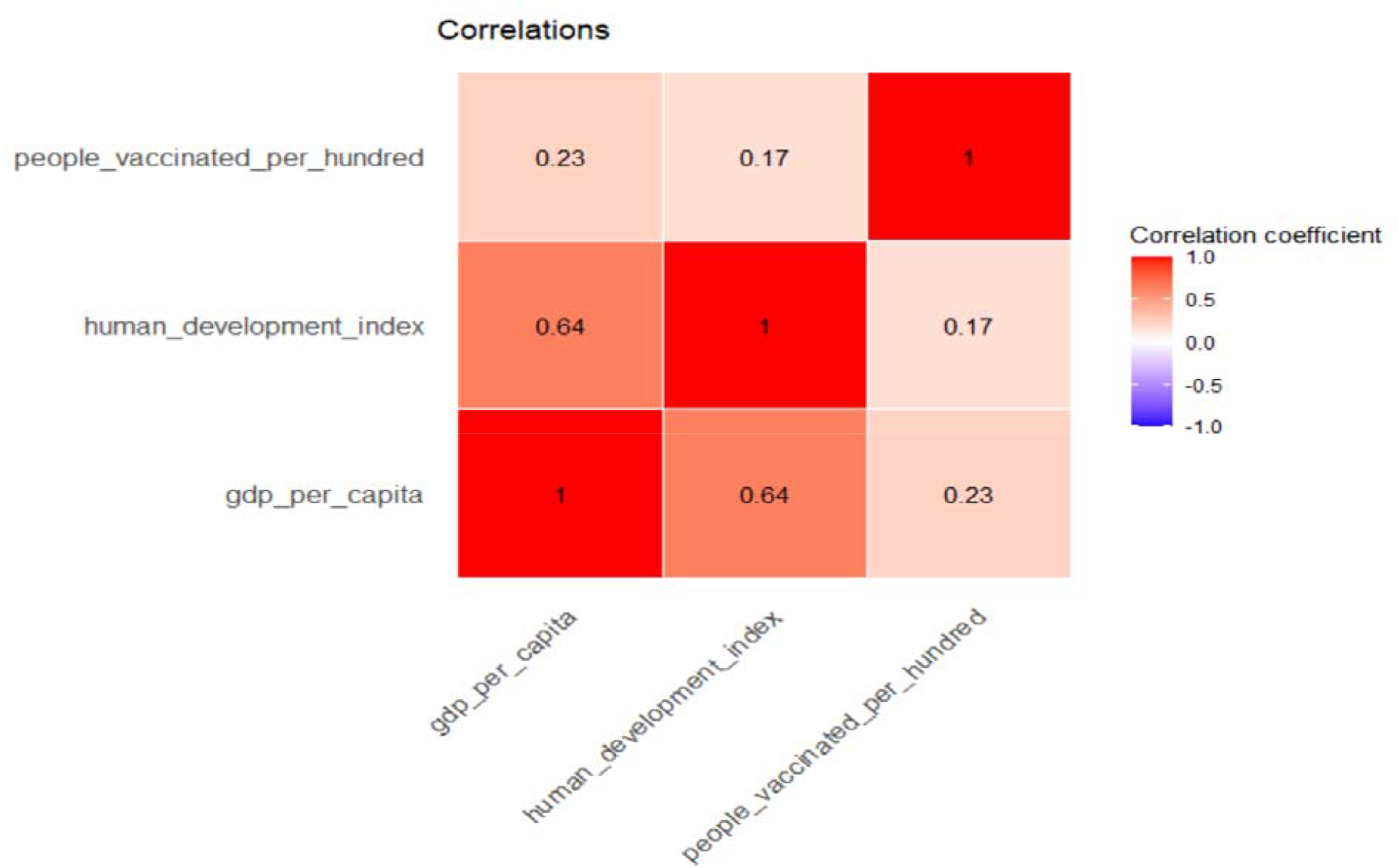

**Figure 2.**
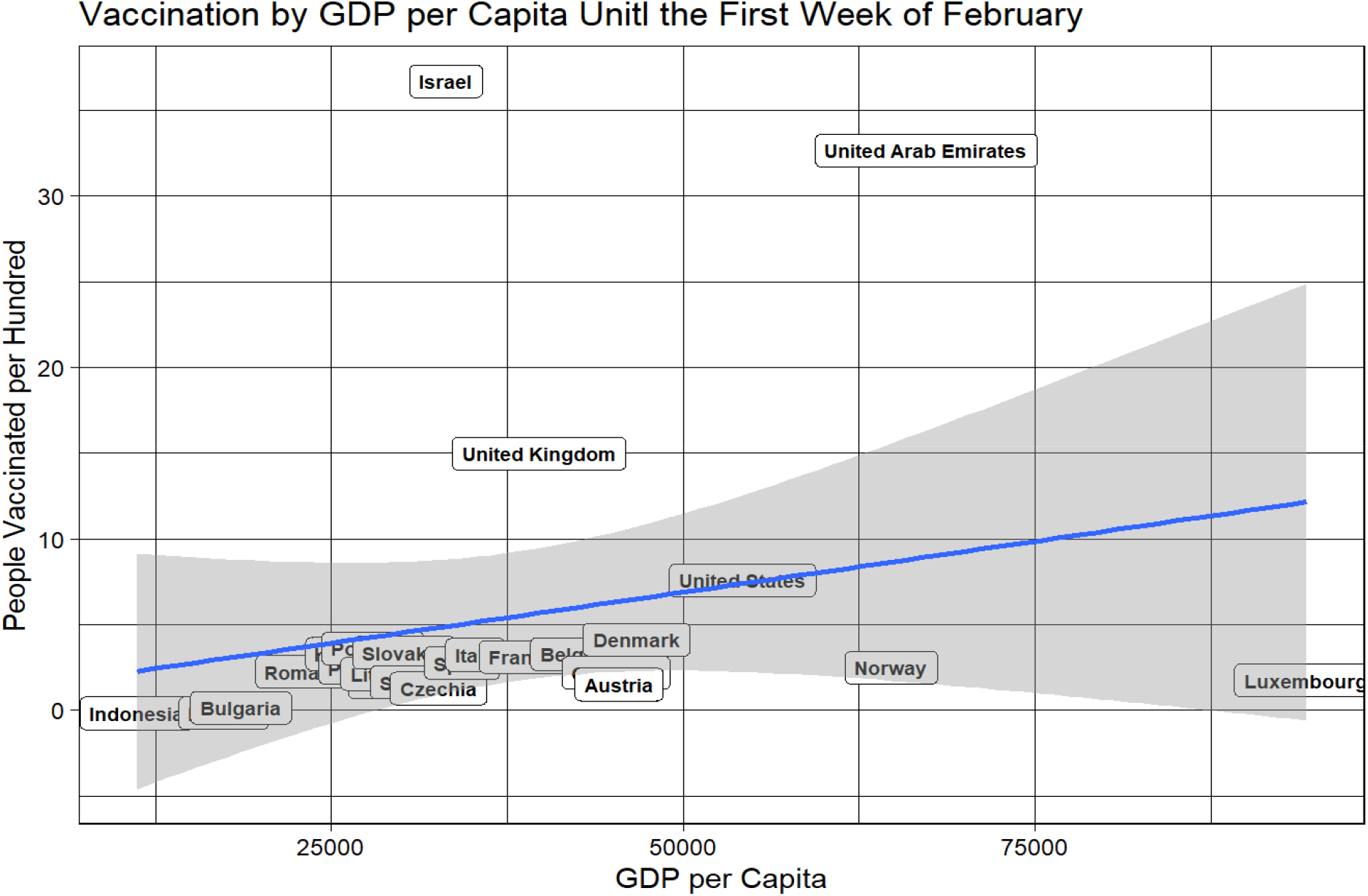

Figure 3 shows the association between HDI and people vaccinated per hundred, which shows that almost all countries have the same performance based on HD.

**Figure 3.**
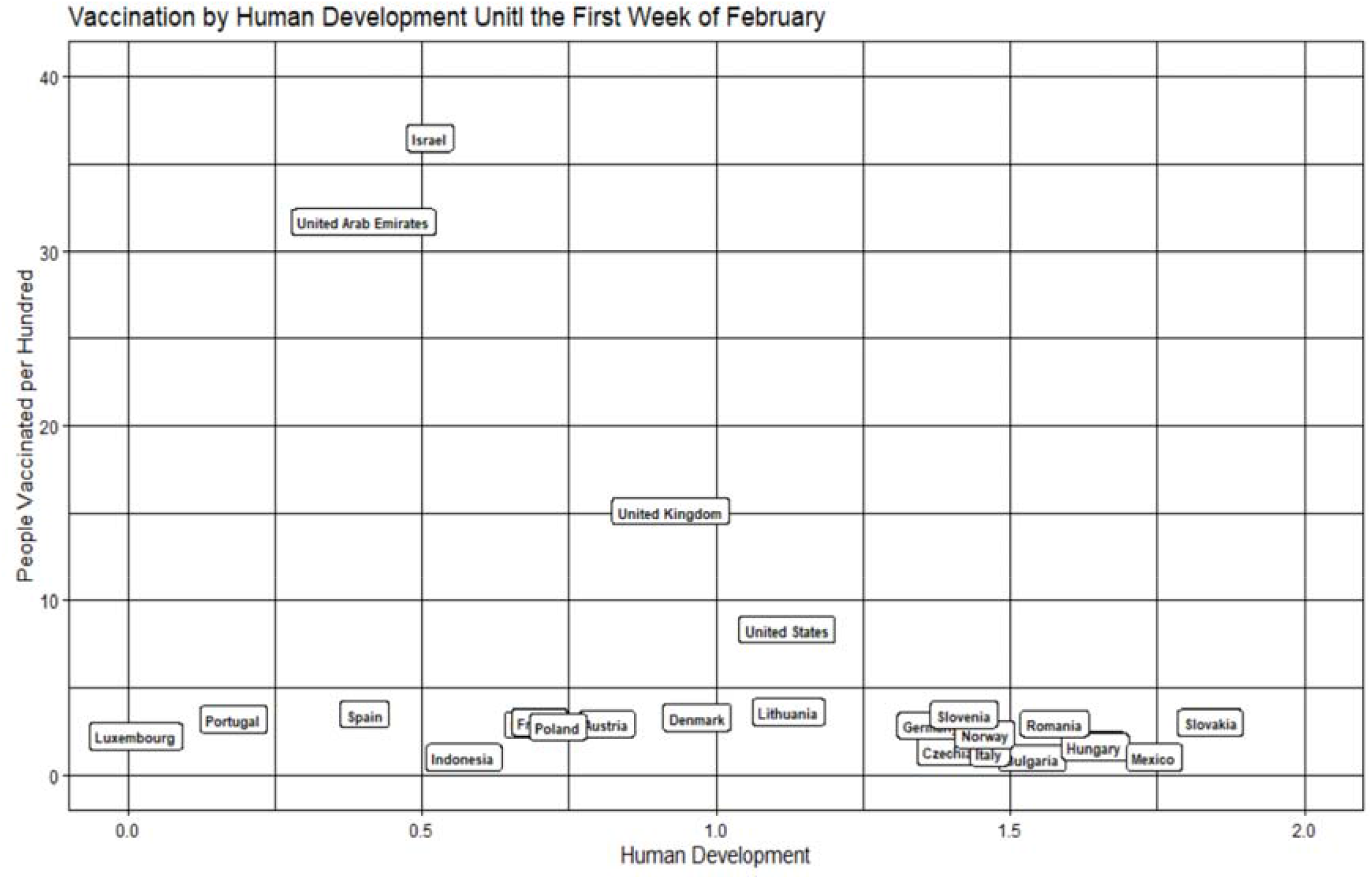

The regression analysis (Table 2) shows higher GDP per Capita is significantly associated with higher vaccine distribution. In the second model, including population density and hospital beds, the odds ratio decreases, but countries with higher GDP per Capita are more likely to have higher vaccination distribution. There is no association between HDI and vaccine distribution, however, higher hospital beds can predict a higher percentage of the population vaccination slightly.

**Table 2.** Regression Analysis (Outcome: People Vaccinated per Hundred)

|  | Odds Ratio (p value) | Odds Ratio (p value) |
| --- | --- | --- |
|  | Model 1 | Model 2 |
| GDP per Capita | 1.38 ** | 1.33 * |
| Human Development Index | 1.23 | 1.06 |
| Hospital bed |  | 1.13* |
| Population density |  | 1.13 |

OR: Odds Ratios; Significant at \* $p \leq 0.05$ . \*\* $p \leq 0.01$ . \*\*\* $p \leq 0.001$

## Discussion

This study shows that higher GDP per capita is significantly associated with greater numbers of vaccinations; however, there was no significant relationship between HDI and vaccine distribution. These differences can be related to their health policies, priorities, and other medical interventions that the mechanisms of this study cannot explain these differences. Our finding shows that Norway with high GDP per capita has a low vaccination distribution, and this low percentage can be relayed to other concerns than medical facilities. A study by Torjesen (2021) shows doctors in Norway were told to evaluate severely frail old patients after receiving the Pfizer vaccine against covid-19, following the deaths of 23 cases shortly after taking the vaccine (7).

Our findings imply that GDP may be more crucial than the HDI in a critical moment such as the COVID-19 pandemic. The reasoning would be that GDP may support the production, test, and distribution greater than HDI, which is a social determinant measure. We can conclude that with a longer duration of time, we may have a more comprehensive assessment between HDI and vaccine distribution,

Although this study shows important implications regarding the relationship between economic, social determinants and vaccine distribution, it has some limitations. First, because of the limitations in the data, we had to restrict the numbers of countries. Also, most of the countries have started vaccinations in less than a month. Therefore, a longer period can help us have a more accurate understanding of factors that influence vaccine distribution. This data was limited to a few economic, health, and social determinants. Information about labs, vaccine producers, and their relationship with these countries could enrich the data.

## Conclusion

Overall, this study has a vital contribution to the COVID-19 research by focusing on GDP’s role. GDP can play a vital role in people’s immunization; however, some countries with a lower GDP per capita still have a better performance. Future studies should focus on the hidden factors that explain what elements can speed up vaccination.

## Data Availability

https://ourworldindata.org/coronavirus

## Code Availability

The code to perform all analyses described in Methods and Usage Notes sections was written in R-3.6.2 and is publicly available at https://ourworldindata.org/coronavirus

## Author Approval

All authors have seen and approved the manuscript.

## Author Contributions

Ali Roghani designed the study and implemented methods and analyses. Samin Panahi contributed to evaluate and develop the methods, and structure. All authors were involved in developing the ideas and drafting the paper.

